## Supplemental Note 1 for "Multicountry genomic analysis underscores regional cholera spread in Africa"

### **SUPPLEMENTARY NOTE 1**

**CholGEN Consortium Authors**

Catholic University of Bukavu. Patrick Musole Bugeme

CDC Malawi. Elizabeth Kampira

Epicentre. Flavio Finger, Rachel Mahamba

Institut National de Recherche Biomédicale (DRC). Joel Kakwanda Kanyama, Emmanuel Lofiko Lokilo, Pauline-Chloé Kayembe Muswamba, Princesse Paku Tshambu

London School of Hygiene and Tropical Medicine. Chloe Hutchins, Jackie Knee

National Health Laboratory & Diagnostic Services (NHLDS/CPHL), Ministry of Health (Uganda): Grace Alenyo, Lydia Bulyaba, Benedict Kanamwanji, Stephen Kanyerezi, Moses Murungi, Ritah Namusoosa, Hellen Rosette Oundo, Joseph Sekate, William Senyonga, Julius Sseruyange, Godwin Tusabe, Tenywa Wilson

National Laboratory of Lubumbashi (DRC): Jacques Muzinga

National Program of Elimination of cholera, Ministry of Health (DRC): Doudou Boloweti Batumbo

National Public Health Laboratory, Ministry of Public Health (Cameroon): Yves Amang, Chanceline Bilounga Ndongo, Yvette Ebogo, Elodie Edinga, Linda Esso, Francoise Fouda, Etienne Guenou, Clarisse Kila, Nadia Jacqueline Mandeng, Moise Christian Junior Meka, Marie Claire Okomo Assoumou, Yvette Wirba, Sylvain Engamba

Nigeria Centre for Disease Control and Prevention: Sophiyah Damilola Adelakun, Osaoghomwen Amiebenomo, James Avong, Deborah Effiong, Eme Ekeng, Kenneth Chukwuemeka Ikeata, Khadijah Imam, Ikechukwu Nnaji, Zayyanatu Nuru, Richard Olulowo Ojedele, Deborah Okomayin, Chidiebere Opara, Victor Oripenaiye

Public Health Institute of Malawi: Arthur Baluwa, Ephrone Keddie Banda, Chifundo Banda, Mphatso Katumbule Bukhu, Yollam Chavula, Moses Chitenje, Michael Hauli, Patrick Kalengo, Mphatso Kanjiru, Grace Kusakara, Innocent Malolo, Happy Manda, Christopher Misomali, Annie Mwale, Andrew Mzumara, Bright Odala, Louis Panja

Rodolphe Merieux Institut National de Recherche Biomédicale-Goma (DRC): Tavia Matamu Bodisa, Jules Namugusha Cizungu, Yves Birindwa Hamisi, Pascal Nzoloka Kabuyaya, Adèle Kamaliro Kavira, Faida Kitoga, Brigitte Modrada Madakpa, Jeriel Mufungizi, Michel Ngimba, Zéphanie Kalimuli Paluku, Espérance Tsilabia Tsiwedi

Zambia Cholera Task Force & Centre for Infectious Disease Research Zambia: Caroline C. Chisenga, Michelo Simuyandi

Zambia National Public Health Institute: Priscilla Nkonde Gardner, Muzala Kapin'a, Peter Chibale Mwansa, Nchimunya Siabeenzu
